# Demonstration of a Longitudinal Action Medical Mission (LAMM) Model to Teach Point-of-Care Ultrasound in Resource-Limited Settings

**DOI:** 10.1101/2020.05.08.20095760

**Authors:** Michael Yao, Lauren Uhr, George Daghlian, Junedh M. Amrute, Ramya Deshpande, Benji Mathews, Sanjay A. Patel, Ricardo Henri, Gigi Liu, Kreegan Reierson, Gordon Johnson

## Abstract

**BACKGROUND:** Short-term medical missions prevail as the most common form of international medical volunteerism, but they are ill-suited for medical education and training local providers in resource-limited settings.

**OBJECTIVE:** The purpose of this study is to evaluate the effectiveness of a longitudinal educational program in training clinicians how to perform point-of-care ultrasound (POCUS) in resource-limited clinics.

**DESIGN:** A retrospective study of such a four-month POCUS training program was conducted with clinicians from a rural hospital in Haiti. The model included one-on-one, in-person POCUS teaching sessions by volunteer instructors from the United States and Europe. The Haitian trainees were assessed at the start of the program and at its conclusion by a direct objective structured clinical examination (OSCE), administered by the visiting instructors, with similar pre- and post-program ultrasound competency assessments.

**RESULTS:** Post-intervention, a significant improvement was observed (*p* < 0.0001), and each trainee showed significant overall improvement in POCUS competency independent of the initial competency pre-training (*p* < 0.005). There was a statistically significant improvement in POCUS application for five of the six medically relevant assessment categories tested.

**CONCLUSION:** Our results provide a proof-of-concept for the longitudinal education-centered healthcare delivery framework in a resource-limited setting. Our longitudinal model provides local healthcare providers the skills to detect and diagnose significant pathologies, thereby reducing avoidable morbidity and mortality at little or no addition cost or risk to the patient. Furthermore, training local physicians obviates the need for frequent volunteering trips, saving costs in healthcare training and delivery.

## Background

Point-of-care medicine is a rapidly emerging field in diagnostic imaging that allows clinicians to administer acute care at the bedside in emergent situations [1]. Namely, point-of-care ultrasound (POCUS) is a reliable diagnostic tool that affords clinicians the ability to examine patients in real time and make clinical assessments without the expense, delay, or radiation associated with other imaging modalities [2]. Given its profound versatility, usability, and low training requirements, POCUS is emerging as an increasingly important tool for bedside diagnosis [3-9].

With reduced costs and a user-friendly portable interface, POCUS holds promise for application in resource-limited settings that often suffer from a lack of access to diagnostic imaging technologies [13]. Previous studies have shown POCUS to be effective in developing countries. For instance, Shah et al. showed that in two district rural hospitals the utilization of ultrasound changed the course of patient care up to 43% of the time [14], while Kotlyar et al. reported a ultrasound-driven change in patient management in 62% of cases in a tertiary care center in Liberia [15]. Furthermore, POCUS education was shown to improve patient care by Mathews et. al., who developed a standardized protocol of POCUS education that resulted in long-term retention of ultrasound image acquisition and interpretation skills [16].

Given the well-documented benefits of POCUS, applying this technology to resource-limited areas would be opportune. Traditionally, international volunteering trips to resource-limited settings are centered around ‘short-term medical missions’ (STMMs), which are largely considered to be cost-ineffective and limited in timespan [17]. One primary driver of this low efficacy is that many pathologies, especially in resource-limited areas, cannot be detected and/or treated in the duration of a single medical mission. Most medical interventions in these settings often require delivery of delayed lab results or treatment of complications secondary to initial treatment, which precludes clinical follow-up especially in the timeframes of typical STMMs [17]. As a result, the existing medical infrastructure is left to bear the consequences of treating an acute influx of patients with already limited resources. Given these drawbacks, follow-up post-STMM treatment may potentially adversely affect the existing patient population as a result of a diversion and dilution of limited medical resources [17]. Specifically, the use of POCUS in medical volunteering trips requires additional training that many clinicians practicing in low-resource medical institutions lack, making incorporating POCUS into medical practice challenging.

In this study, we propose a novel educational model, ‘longitudinal action medical missions’ (LAMMs), and discuss how this model transcends many of the limitations faced by traditional STMMs. The LAMM model provides a unique framework for sustained medical education while minimizing costs and burden on the instructors. This was accomplished through an extended program that emphasizes hands-on training at the bedside, as opposed to the traditional patient care-oriented model of STMMs. Traditional short courses can get trainees familiar with POCUS, but their skills can often decay overtime as a result of a lack of longitudinal repetition. Alternatively, trainees are more likely to retain skills and continue utilizing POCUS as a part of their standard practice with a longitudinal training program over months, rather than an intensive, traditional two- or four-day course. However, many volunteer POCUS experts are unable to dedicate many months away from their respective practices to teach such longitudinal courses.

To address this limitation, we created multiple teams of two to three instructors where each team visits the low-resource medical clinic for one week, spread out over a four-month period. This provided continuous mentorship for the POCUS trainees over the course of many months and allows for longitudinal hands-on training and evaluation. Furthermore, utilizing an array of instructors from Europe and the United States provides a broad perspective in teaching POCUS. The objective of the bedside training sessions is to work with the trainees individually and teach them fundamental POCUS skills through real-time clinical scenarios. Training local physicians reduces the need for frequent volunteering trips in the long run and has the potential to save tremendous costs in healthcare delivery. We show that by equipping and training Haitian physicians with state-of-the art diagnostic tools, such as POCUS, we can correctly diagnose many prevalent pathologies that often go misdiagnosed due to a lack of training and resources.

## Methods

We retrospectively evaluated the efficacy of the proposed LAMM model implemented over the span of four months. Study participants were clinicians from the 68-bed Alma Mater Hospital in Gros-Morne, Haiti, a small rural community on the north side of the Caribbean island. The program consisted of 12 local Haitian physicians, with 4 of the 12 completing the full 4-month training program while the remaining 8 engaged in a fraction of the training program. Our POCUS educators were practicing physicians across a wide variety of specialties including radiology, internal medicine, family practice, and emergency medicine.

### Program Description

The goal of our hospital-based POCUS training program was to demonstrate a proof-of-concept methodology in incorporating the usage of POCUS in resource-limited settings to ultimately improve the accuracy and timeliness of diagnoses and safety of procedures by utilizing ultrasound guidance. Our POCUS training program consisted of nine weeks of in-person training sessions spread over the course of four months. There were two week-long sessions for a general introduction to ultrasound, followed by seven weeks of POCUS training with one- to two- week separation periods interspersed throughout the training program. The overall training program consisted of a total of six ultrasound fundamentals: neck, lung, abdomen, cardiac, lower extremity, and soft tissue (**Table 1**). The curriculum was tailored to the individual specialties of the volunteer POCUS instructors on each week-long trip; collectively, they were able to cover a variety of hands-on concepts, as detailed in **Table 1**. Our pre- and post-training program objective structured clinical examination (OSCE) assessments were designed to assess the basics of POCUS, and were adapted from the Comprehensive Hospitalist Assessment and Mentorship with Portfolios (CHAMP) ultrasound program from Mathews et al [16]. All volunteer instructors followed a standard set of ultrasound education principles to guide the in-person training of participants.

**Table 1:** Fundamentals of the ultrasound assessment.

| **Examination:** | **Fundamentals of Identifying and Interpreting POCUS Images:** |
| --- | --- |
| Neck ultrasonography | Carotid artery in transverse and longitudinal planes<br>Color Doppler of carotid artery<br>Internal jugular vein in transverse and longitudinal planes |
| Lung ultrasonography | Pleural line (lung sliding and A lines)<br>Presence of pleural effusion<br>Costophrenic recess |
| Abdominal ultrasonography | Liver and hepatorenal space<br>Spleen and splenorenal space<br>Kidneys and bladder in transverse and longitudinal planes<br>Abdominal aorta in transverse and longitudinal planes |
| Echocardiography | LV, RV, LA, aorta, mitral and aortic valves, and pericardium in parasternal long axis<br>Papillary muscles in parasternal short axis<br>Four chambers and valves in the apical four-chamber view |
|  | Hepatic vein and IVC in longitudinal axis with IVC entry into RA in subcostal view |
| Lower extremity ultrasonography | Femoral vein and artery with compression<br>Greater saphenous and popliteal vein with compression |
| Musculoskeletal soft tissue ultrasonography | Achilles tendon<br>Skin<br>Muscle |

### Online Modules and Supplemental Reading

Online training modules were provided by SonoSim, Inc. and featured a handheld probe simulator for interactive feedback. We asked the study participants to review the relevant online modules prior to the weekly group sessions that focused on a particular POCUS topic. For example, the participants were asked to review the echocardiography online module from SonoSim, Inc. prior to a week-long, in-person didactic sessions with the echocardiography specialist.

This online module-based training was supplemented with *Point-of-Care Ultrasound* POCUS-training textbook from Soni et al, which also acted as a reference guide for the study participants [18].

### Bedside POCUS Training

As aforementioned, the core of our POCUS training program involved a series of nine week-long, hands-on training sessions that together formed a single LAMM. The program utilized the Philips Lumify portable ultrasound system consisting of their S4-1 cardiac, C5-2 curvilinear, and L12-4 linear transducers. The Lumify system remained on-site indefinitely for continued usage by the trainees after the conclusion of the training program.

### Reacts-enabled Tele-ultrasound Training and Collaboration

In addition to the in-person training provided by volunteer POCUS experts, we used the Lumify with Reacts (Remote Education, Augmented Communication, Training and Supervision) integrated tele-ultrasound software developed in collaboration by Philips ultrasound and Innovative Imaging Technologies (IIT), Inc. in order to work with the study participants remotely in real-time. Reacts is a collaborative platform developed by IIT that allows for real-time remote medical education and consultation. The hand-held ultrasound system simultaneously streams ultrasound images along with video and audio in real-time. The system was used to continue ultrasound education during the limited number of weeks where POCUS educators were not available to travel to Haiti. These remote training sessions were conducted approximately three times over the course of the four-month data collection period using a reliable cellular or Wi-Fi connection.

### Assessments

Trainees were assessed in six core areas of ultrasound in terms of imaging and interpretation: neck, lung, abdomen, cardiac, lower extremity, and soft tissue. In each of these areas of assessment, there were multiple skills and participants were evaluated on a scale of 0 to 2 for each skill. A score of 0 represents the inability to generate the ultrasound image, 1 represents the ability to generate the ultrasound image but an inaccurate interpretation of the image, and 2 represents the ability to both generate and interpret the ultrasound image. This ultrasound skills assessment was adapted from the CHAMP ultrasound program [16].

### Statistical Analysis

Assessment scores per core area with multiple skills were reported as the average of values, with the minimum possible score being 0 and the maximum being 2. Comparisons of assessment scores were performed using paired-samples t-test. Differences were considered statistically significant when p-value was less than 0.05. Statistical Package for the Social Sciences (SPSS) (IBM SPSS Statistics for Mac, Version 26.0. Armonk, NY: IBM Corp) was used for analysis.

## Results

The assessment score data for our sample of *n =* 4 physicians that completed the training program is shown in **Fig. 1**. The mean score across all assessments in the fundamental areas of POCUS are shown in **Fig. 1(a)**; prior to our program, the average score per assessment was 0.47, which improved to 1.68 (*p* < 0.0001) after the training program. Furthermore, each physician showed substantial overall improvement in POCUS competency (**Fig. 1(b)**), independent of the initial competency pre-training (p < 0.004). Finally, there was an overall improvement in POCUS application for five of the six medically relevant assessment categories tested: neck, lung, abdomen, cardiac, and lower extremity imaging (**Fig. 1(c)**).

**Figure 1.**
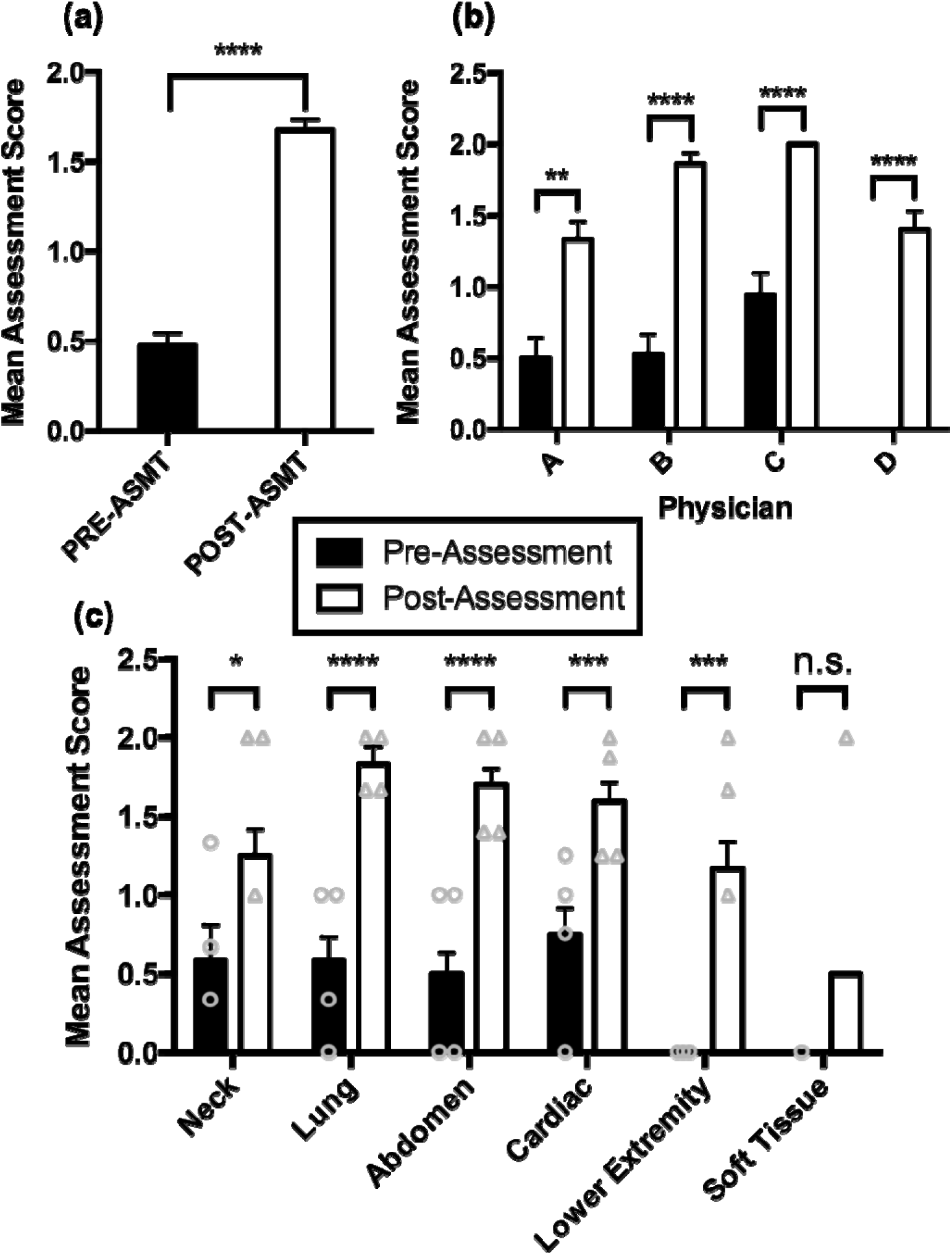
Mean assessment score of physicians pre- and post-training program with 95% confidence level. **(a)** The average POCUS competency score per assessment (trainees were given a 0, 1, or 2) pre- and post-training. Paired *t*-test, df=66, *p <* 0.0001. **(b)** The average POCUS competency score per assessment pre- and post-training for each physician that completed the entirety of the training program. Paired *t*-test, df=14, 17, 18, and 14 for Physicians A, B, C, and D, respectively, *p* = 0.0032 for Physician A, *p* < 0.0001 for Physicians B - D. **(c)** The average POCUS competency score per assessment pre- and post-training for each of the six medically relevant categories, as per the delineated categories in our assessment. Paired sample *t*-test, df=8, 11, 19, 15, and 8 for neck, lung, abdomen, cardiac, and lower extremity/DVT, respectively. *p* = 0.0207, *p* < 0.0001, *p* < 0.0001, *p* = 0.0002, and *p* < 0.0001, for neck, lung, abdomen, cardiac, lower extremity/DVT, respectively. Due to limited data in soft tissue category, no statistical testing could be conducted in this category. Vertical lines above each bar indicate one standard error of the mean on either side of the mean; absence of a bar indicates a mean of 0, and absence of a line above a bar indicates no reportable standard error. PRE-ASMT = Pre-assessment; POST-ASMT = Post-assessment.

## Discussion

The LAMM model demonstrates a statistically significant trend of improvement in most categories of assessment. Given that our sample population consisted of Haitian physicians, the baseline assessment scores shown in **Fig. 1(b)** is expected, as the Haitian medical education covers many of the fundamentals of neck, lung, abdomen, and cardiac anatomy. In contrast, trainees exhibited no prior knowledge regarding image acquisition and interpretation with respect to lower extremities/DVTs and soft tissue ultrasonography, which are often considered to be more advanced ultrasound techniques. Improvements in assessment scores were observed in all categories of ultrasound image acquisition and interpretation regardless of previous experience.

One limitation to this study was the trainee dropout rate. The dropout of eight out of twelve clinicians could have led to attrition bias in the assessment scores. Compared to other ultrasound training programs reported in the literature [19, 20], our dropout rate is expected. Follow-up with the clinicians who did not complete the program would be informative to understand the attitudinal, psychomotor-skill related, cultural, language, and/or clinical barriers to learning POCUS in a resource-limited setting. Despite this dropout rate, the longitudinal nature of this ultrasound training program resulted in highly skilled POCUS trainees, a few of which extended their clinical employment with the Alma Mater Hospital to continue their POCUS training of the remaining clinicians, including some that did not finish the training program described herein. These new POCUS-trained clinicians serve as evidence for the efficacy of the longitudinal model over the traditional short-term trips—by continually training local physicians, they are reducing the need for future volunteer trips.

Qualitatively, Physician C in **Fig. 1(b)** reported to have studied textbook ultrasound training material prior to the beginning of the program, while the other physicians reported no such self-studying [18]. The data suggests that self-study, even without hands-on practice, could be beneficial for improved assessment scores both during the pre-study assessment and post-study assessment. Indeed, among all the study participants that completed this training program, Physician C performed the best in both the pre-training and post-training assessments conducted. Further randomized studies are needed with a larger number of study participants in order to better investigate if pre-program studying is beneficial to physician competency both during and following the program.

During this study, remote tele-ultrasound education was explored using the Reacts software integrated into the Philips Lumify ultrasound system. We found the Lumify with Reacts tele-ultrasound to be user-friendly and convenient even in this resource-limited setting. This suggests that Lumify with Reacts tele-ultrasound is a viable option for continued education and training even in resource-limited settings. In general, Lumify with the Reacts integration used in this study presents an exemplifying instance of the growing field of healthtech advancements, enabling long-term cost reductions and the ability to pursue sustainable LAMM models. This platform has the potential to empower local POCUS leaders to scale up training of other nearby communities with the support and expertise of international volunteer instructors, circumventing language/cultural barriers and high transportation costs, and reducing the target communities’ dependence on foreign involvement in the long run. This concept of self-sustainability is a particularly promising feature of the LAMM model, as evidenced by the trainees training additional Haitian clinicians.

We envision the potential for many long-term benefits of pursuing LAMMs as opposed to STMMs in terms of cost, sustainability, and self-reliability. As discussed previously, the primary limitation with most STMMs is that they are unable to build on previous work done with the communities of interest; every medical mission is essentially conducted independently of adjacent ones. Thus, while current STMMs can provide temporary relief with varying efficacy, they are not appropriately structured to provide medical assistance in the form of specialized medical education, technical training, and other forms of aid that cannot feasibly be accomplished within the mission’s short time frame. The LAMM model is a feasible solution to address this problem, and we have demonstrated its use in bringing POCUS education and technology to an under-resourced setting.

## Conclusions

The proposed LAMM model successfully demonstrates a pragmatic method of incorporating POCUS into an existing medical infrastructure in a resource-limited setting. Our POCUS training program significantly improved the clinician study participants’ ultrasound acquisition skills, and thereby patient care. By training local physicians through a longitudinal model and equipping them with state-of-the-art technology, we transcend many of the limitations suffered by short-term volunteering projects and promote self-sustainability. This eliminates the need for on-going volunteering trips thereby saving healthcare costs. Additional technological advancements, such as real-time tele-ultrasound training platforms and lower-cost ultrasound transducers will synergistically allow for more effective changes to community healthcare systems and improvements to differential diagnoses and patient treatments over the long term.

## Data Availability

The data that support the findings of this study are available from the first author upon reasonable request.

## Acknowledgments

The authors would like to thank the volunteer doctors that contributed to the project discussed herein, including Karoline Lund, MD; Martin Rofael, MD; Nils Petter Oveland, MD, PhD; Victoria Vatsvag, Karen Cosby, MD; Sarah Moore, MD; Anna Maw, MD; Stefan Tchernodrinski, MD; and Danielson Jesper, MD. We also thank the Philips Point-of-Care Ultrasound Business and Philips representative Toni Burkett for their extensive support of this study, and for providing the Lumify with Reacts ultrasound system used in the training sessions discussed herein. We also acknowledge the generous assistance made by SonoSim, Inc. and Innovative Imaging Technologies in contributing to the success of this project. We also thank Sling Health, Los Angeles Chapter and general Sling Health Network for their support, in addition to the individuals at Alma Mater Hospital in Gros-Morne, Haiti for working with us in this study. Finally, we thank Dr. Nilam Soni and Elsevier Publishing for the donation of their POCUS textbooks for each of our trainees.

## Ethics Approval

This study was deemed exempt and approved by the Legacy Emanuel Institutional Review Board (IRB) (Portland, OR) under exempt certification for research conducted in established or commonly accepted educational settings, involving normal educational practices.

## References

1. Rooney KD, Schilling, UM. Point-of-care testing in the overcrowded emergency department–can it make a difference? Crit Care 2014; 18(6):692.

2. Greaves K, Jeetley P, Hickman M, et al. The use of hand-carried ultrasound in the hospital setting-A cost-effective analysis. J Am Soc Echocardiogr 2005; 18(6):620–625.

3. Filly RA. Ultrasound: the stethoscope of the future, alas. Radiology 1988; 167(2):400.

4. Mantuani D, Frazee BW, Fahimi J, et al. Point-of-care multi-organ ultrasound improves diagnostic accuracy in adults presenting to the emergency department with acute dyspnea. West J Emerg Med 2016; 17(1):46.

5. Cortellaro F, Ferrari L, Molteni F, et al. Accuracy of point of care ultrasound to identify the source of infection in septic patients: a prospective study. Intern Emerg Med 2017; 12(3):371–378.

6. Epema AC, Spanjer MJ, Ras L, et al. Point-of-care ultrasound compared with conventional radiographic evaluation in children with suspected distal forearm fractures in the Netherlands: a diagnostic accuracy study. Emerg Med J 2019; 36(10):613–616.

7. Vrablik ME, Snead GR, Minnigan HJ, et al. The diagnostic accuracy of bedside ocular ultrasonography for the diagnosis of retinal detachment: a systematic review and meta-analysis. Ann Emerg Med 2015; 65(2):199–203.

8. Godown J, Lu JC, Beaton A, et al. Handheld echocardiography versus auscultation for detection of rheumatic heart disease. Pediatrics 2015; 135(4):e939-e944.

9. Mok KL. Make it SIMPLE: enhanced shock management by focused cardiac ultrasound. J Intensive Care Med 2016; 4(1):51.

10. Alpert EA, Amit U, Guranda L, et al. Emergency department point-of-care ultrasonography improves time to pericardiocentesis for clinically significant effusions. Clin Exp Emerg Med 2017; 4(3):128.

11. Adhikari S, Stolz L, Amini R, et al. Impact of point-of-care ultrasound on quality of care in clinical practice. Rep Med Imaging 2014; 7: 81–93.

12. Zieleskiewicz L, Muller L, Lakhal K, et al. Point-of-care ultrasound in intensive care units: assessment of 1073 procedures in a multicentric, prospective, observational study. Intensive Care Med 2015; 41(9):1638–1647.

13. Sippel S, Muruganandan K, Levine A, et al. Use of ultrasound in the developing world. Int J Emerg Med 2011; 4(1):72.

14. Shah SP, Epino H, Bukhman G, et al. Impact of the introduction of ultrasound services in a limited resource setting: rural Rwanda 2008. BMC Int Health Hum Rights 2009; 9(1):4.

15. Kotlyar S, Moore CL. Assessing the utility of ultrasound in Liberia. J Emerg Trauma Shock 2008; 1(1):10.

16. Mathews BK, Reierson K, Vuong K, et al. The design and evaluation of the comprehensive hospitalist assessment and mentorship with portfolios (CHAMP) ultrasound program. J Hosp Med 2018; 13(8):544–550.

17. Bauer I. More harm than good? The questionable ethics of medical volunteering and international student placements. Tropical Diseases, Travel Med and Vaccines 2017; 3(1):5.

18. Soni N, Arntfield R, Kory P. Point-of-Care Ultrasound. Elsevier Health Sciences 2014.

19. Henwood PC, Rempell JS, Liteplo AS, et al. Point-of-care ultrasound use over six-month training period in Rwandan district hospitals. Afr J Emerg Med 2013; 3(4):S5-S6.

20. Chung H. Teaching Point-of-Care Ultrasound (POCUS) to Visiting Iraqi Physicians Using the Train-the-Trainer Model. J Glob Health 2018.

